# A model for COVID-19 prediction in Iran based on China parameters

**DOI:** 10.1101/2020.03.19.20038950

**Authors:** Bushra Zareie, Amin Roshani, Mohammad Ali Mansournia, Mohammad Aziz Rasouli, Ghobad Moradi

## Abstract

**Background:** The rapid spread of COVID-19 virus from China to other countries and outbreaks of disease require an epidemiological analysis of the disease in the shortest time and an increased awareness of effective interventions. The purpose of this study was to estimate the COVID-19 epidemic in Iran based on the SIR model. The results of the analysis of the epidemiological data of Iran from January 22 to March 8, 2020 were investigated and the prediction was made until March 29, 2020.

**Methods:** By estimating the three parameters of time-dependent transmission rate, time-dependent recovery rate, and time-dependent mortality rate from Covid-19 outbreak in China, and using the number of Covid-19 infections in Iran, we predicted the number of patients for the next month in Iran. Each of these parameters was estimated using GAM models. All analyses were conducted in R software using the mgcv package.

**Findings:** On average, 925 people with COVID-19 are expected to be infected daily in Iran. The epidemic peaks within one week (15.03.2020 to 03.21.2020) and reaches its highest point on 03.18.2020 with 1126 infected cases.

**Conclusion:** The most important point is to emphasize the timing of the epidemic peak, hospital readiness, government measures and public readiness to reduce social contact.

## Introduction

Transmission of coronavirus-associated pneumonia 2019 (COVID-19) began in Wuhan, China on December 31, 2019 (1). According to the latest WHO report on 11 March 2020, there were 118,329 confirmed cases and 4627 deaths worldwide, with 8042 confirmed cases and 291 deaths reported in Iran (2). The World Health Organization (WHO) named it a global pandemic on 11 March 2020 because of the rapid outbreak of the disease worldwide (3). The course of each epidemic depends on a number of important factors such as the Basic Reproductive Rate, the doubling time or serial interval, and the fatality rate. Estimating the epidemiological factors of COVID-19 and modeling-based prediction are useful in assessing epidemic transmission rates, predicting epidemic trends, and designing control measures (4, 5).

The rapid spread of COVID-19 virus from China to other countries and outbreaks of disease require an epidemiological analysis of the disease in the shortest time and an increased awareness of effective interventions. What happened in China showed that quarantine, evacuation, and isolation of infected populations can become epidemic. This impact of the COVID-19 response in China is encouraging for many countries where COVID-19 has begun to expand. However, it is not clear that other countries can implement the measures that the Chinese eventually adopted to reduce and control the incidence in the future (4, 6).

There are various models for predicting the spread of viruses. Along with vaccines or diagnostic tests, mathematical modeling can be a useful tool for designing strategies for prediction and designing appropriate interventions for rapid control of infectious diseases if not effectively treated. The purpose of this study was to estimate the COVID-19 epidemic in Iran based on the SIR model. The results of the analysis of the epidemiological data of Iran from January 22 to March 8, 2020 were investigated and the prediction was made until March 29, 2020.

### Methods

A simple model of SARS and the precise prediction by Zhang et,al.(7) motivated us to use this model for Covid19. Limited laboratory kits, prolonged test results, people’s lack of awareness and non-referral to medical centers led to incomplete registration of infected, suspected or dead cases in the onset days of the epidemic in Iran. In this situation, it is better to use a model with few parameters and include time-dependent rates. Suppose equation (1) includes three parameters: β(t) time-dependent transmission rate, Y(t) time-dependent recovery rate, μ(t) time-dependent death rate, and time-dependent infected number. The number of patients in our model is a function of time. We predict the number of patients day by day. Therefore, we use the second equation that is based on the first equation.

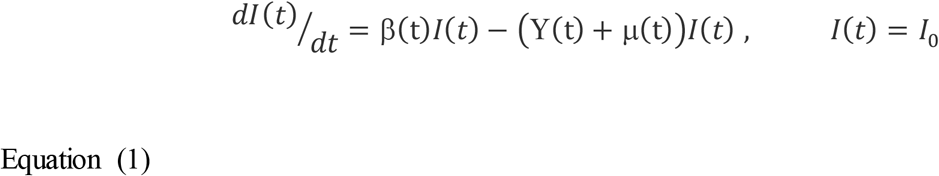

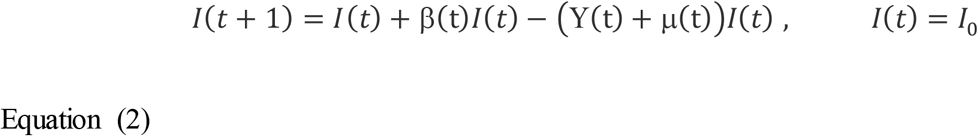

Each parameter is defined as follows:

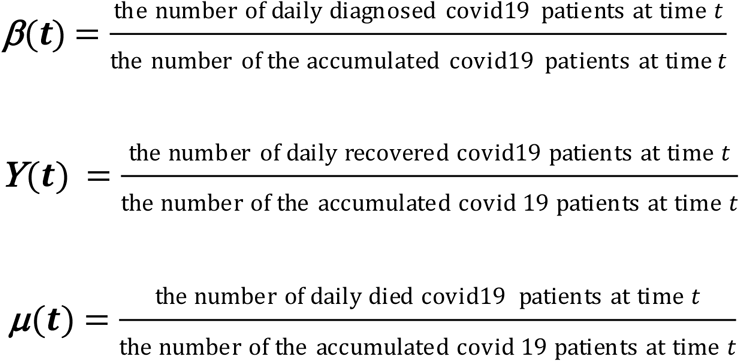

We fit model in three steps.

**Step 1**: In the first step, we analyzed the Chinese data during the outbreak of covid19 and obtained the China model. This allowed us to estimate parameters by accessing more data Figures (1-4).

**Figure 1.**
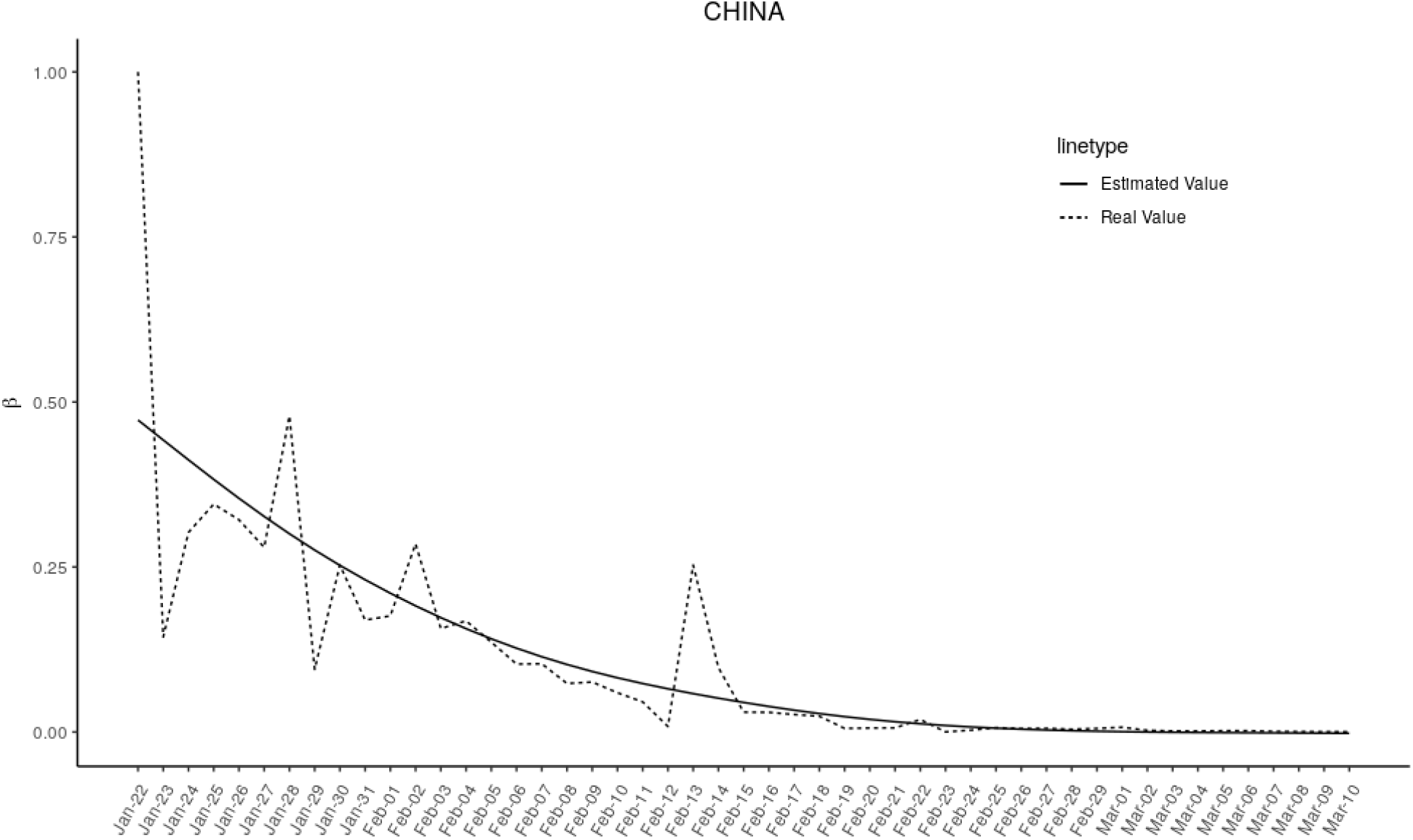
Time-dependent transmission rate people in china from 22.01.2020 to 10.03.2020.

**Figure 2:**
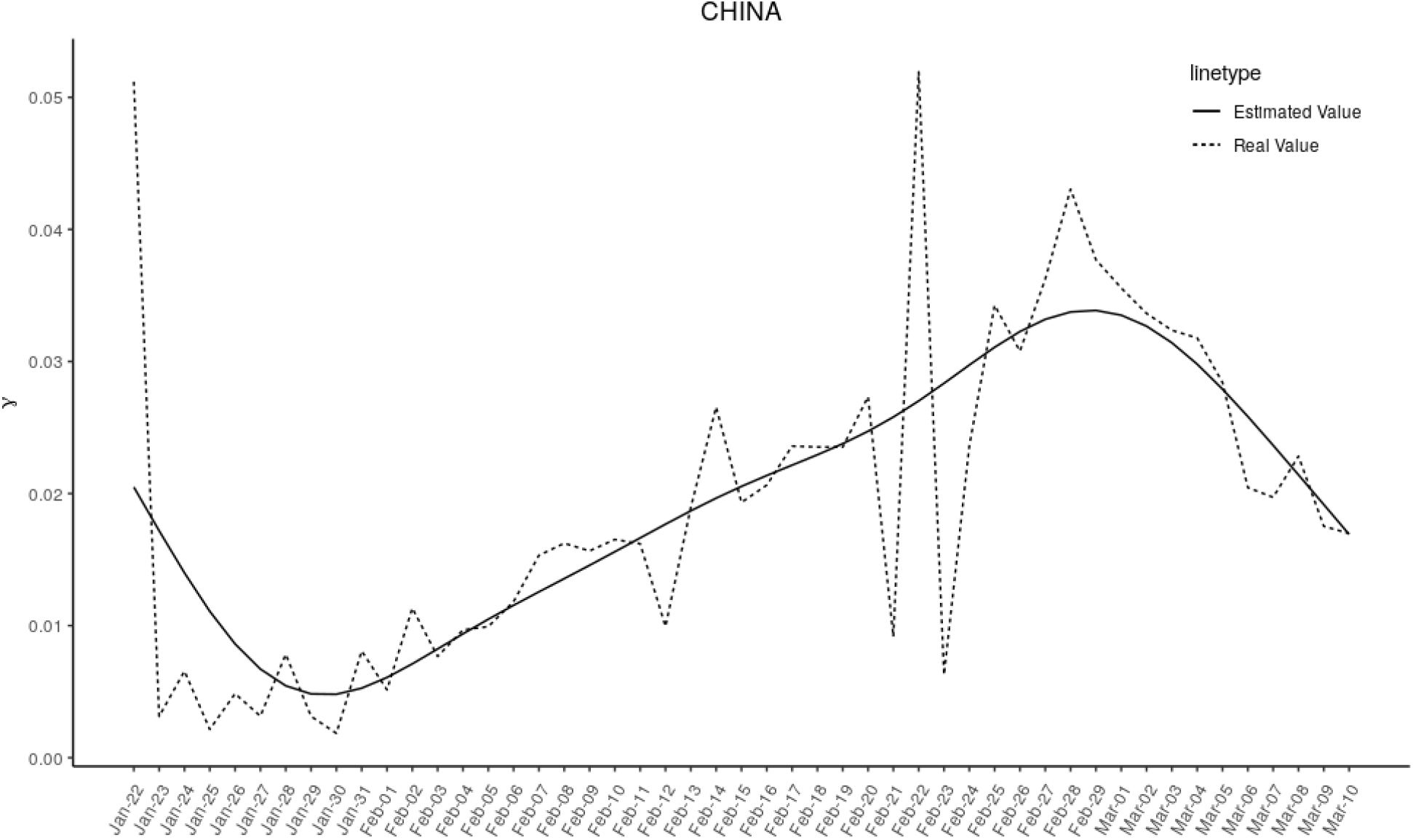
Time-dependent recovery rate people in china from 22.01.2020 to 10.03.2020.

**Figure 3:**
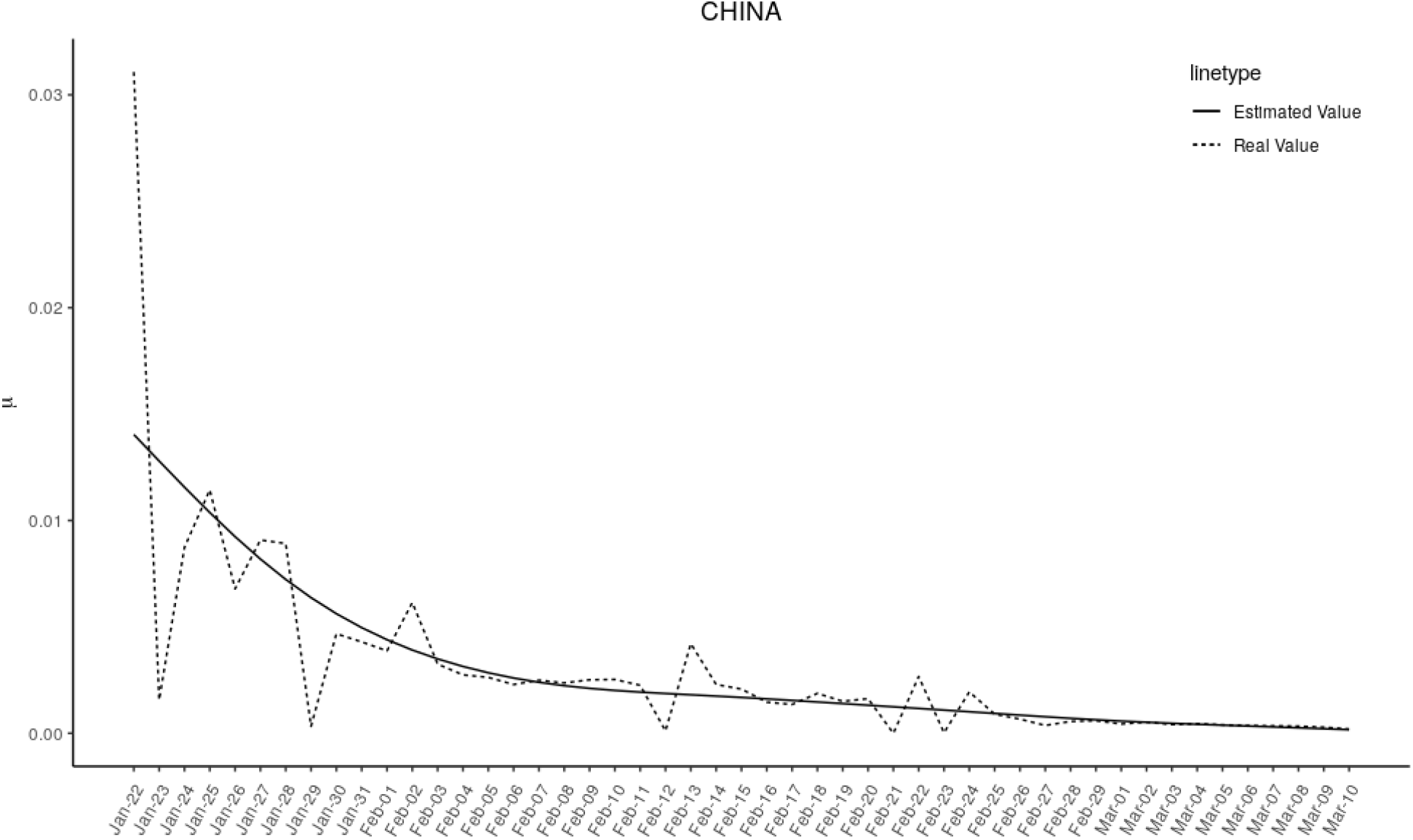
Time-dependent death rate people in china from 22.01.2020 to 10.03.2020.

**Figure 4:**
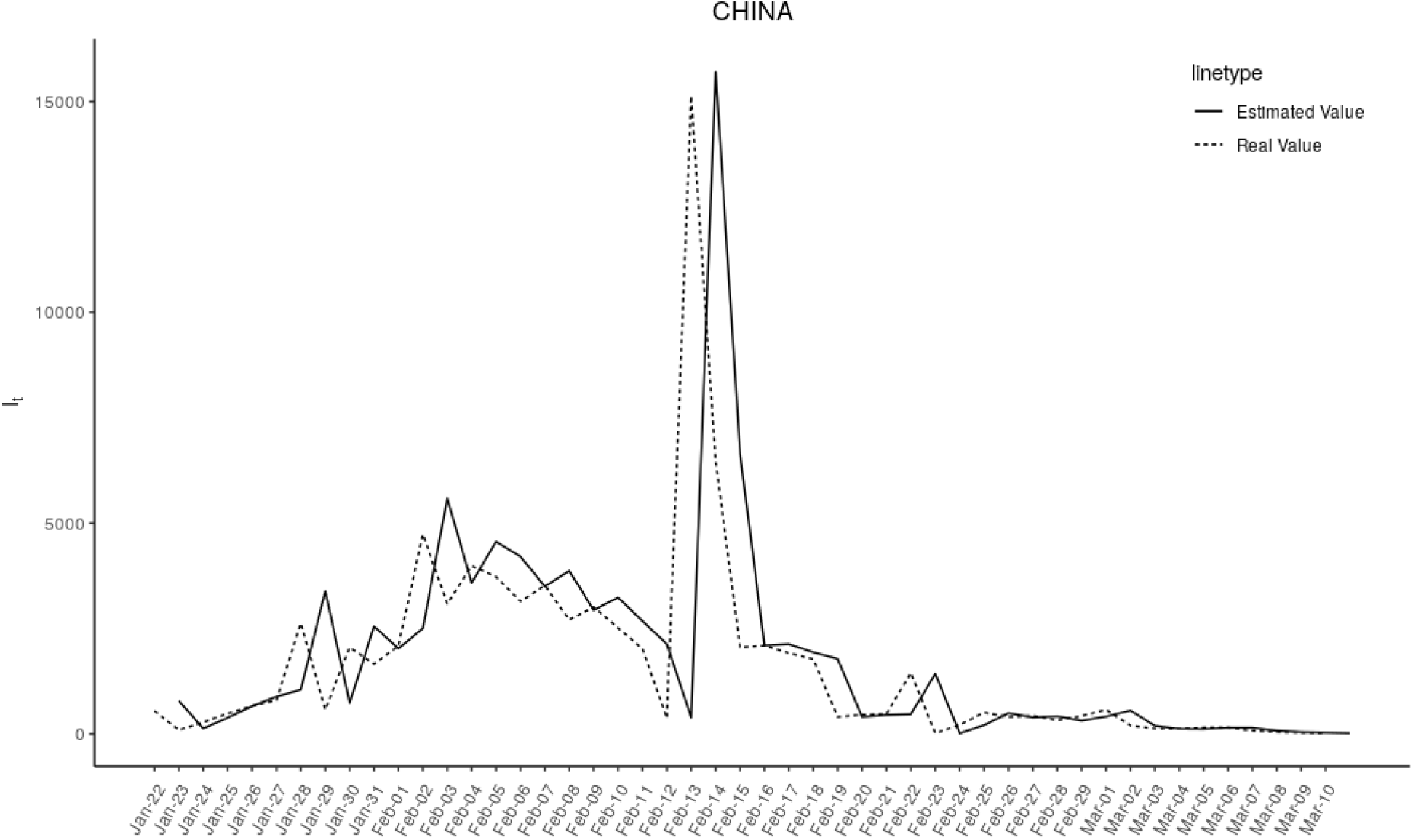
Number of infected people in china from 22.01.2020 to 10.03.2020.

**Step2**: We put the fitting function for β(t),(*t*),(*t*) in the first step along with the actual trend parameters of Iran in Figures (5-7) and found that the behavior parameters of Iran are similar to China.

**Figure 5:**
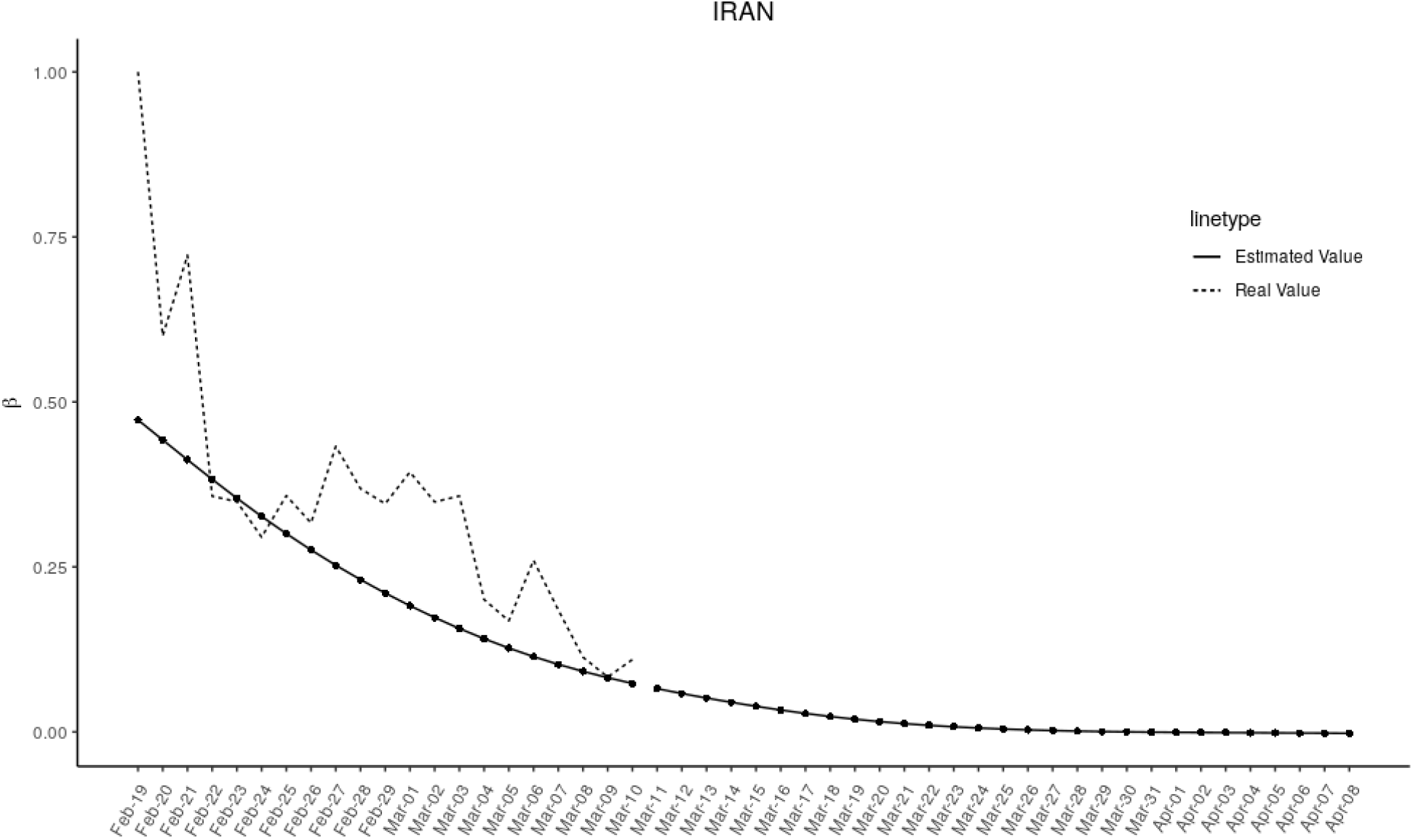
Time-dependent transmission rate people in Iran with β (t) estimated of China.

**Figure 6:**
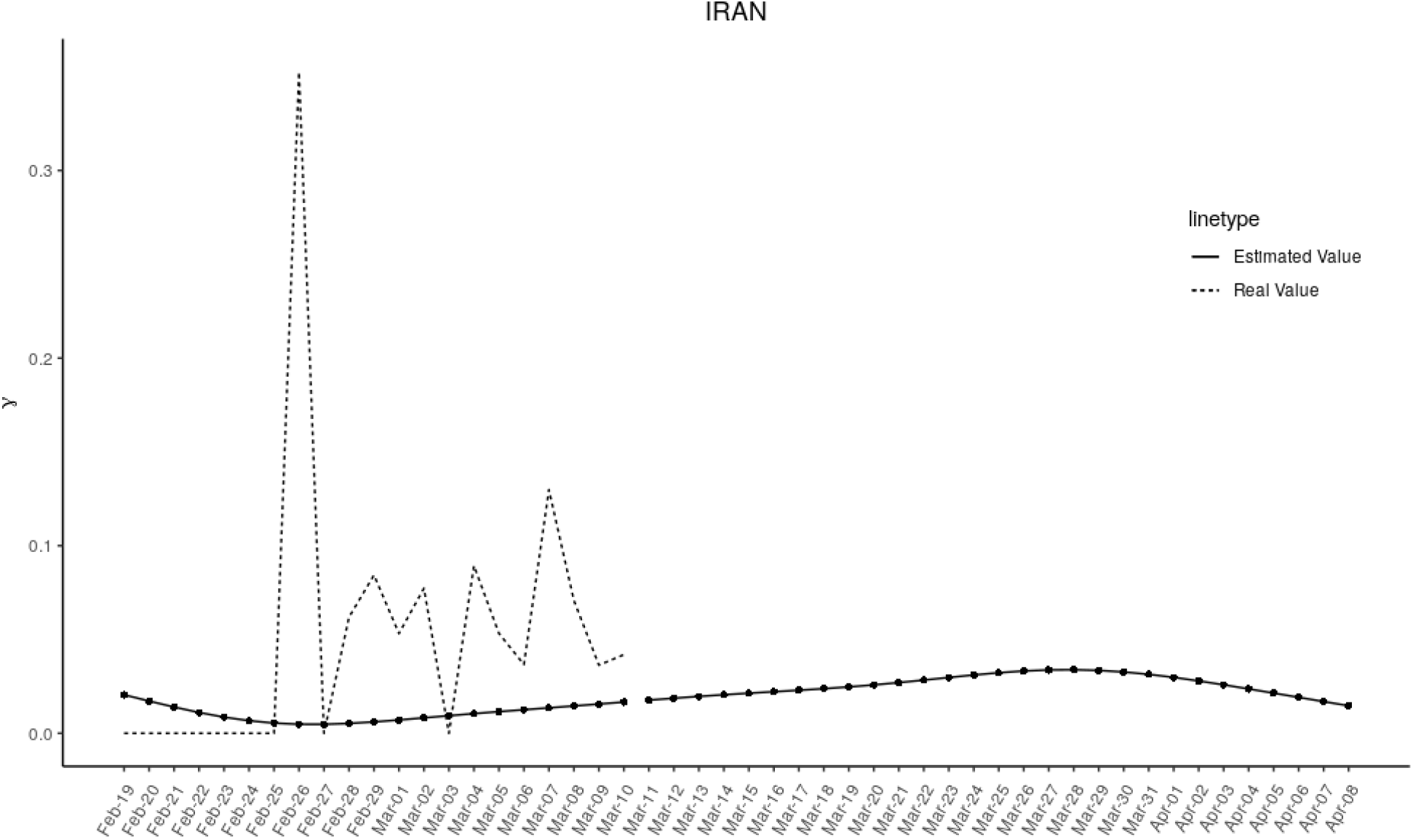
Time-dependent recovery rate people in Iran with Y(t) estimated of China.

**Figure 7:**
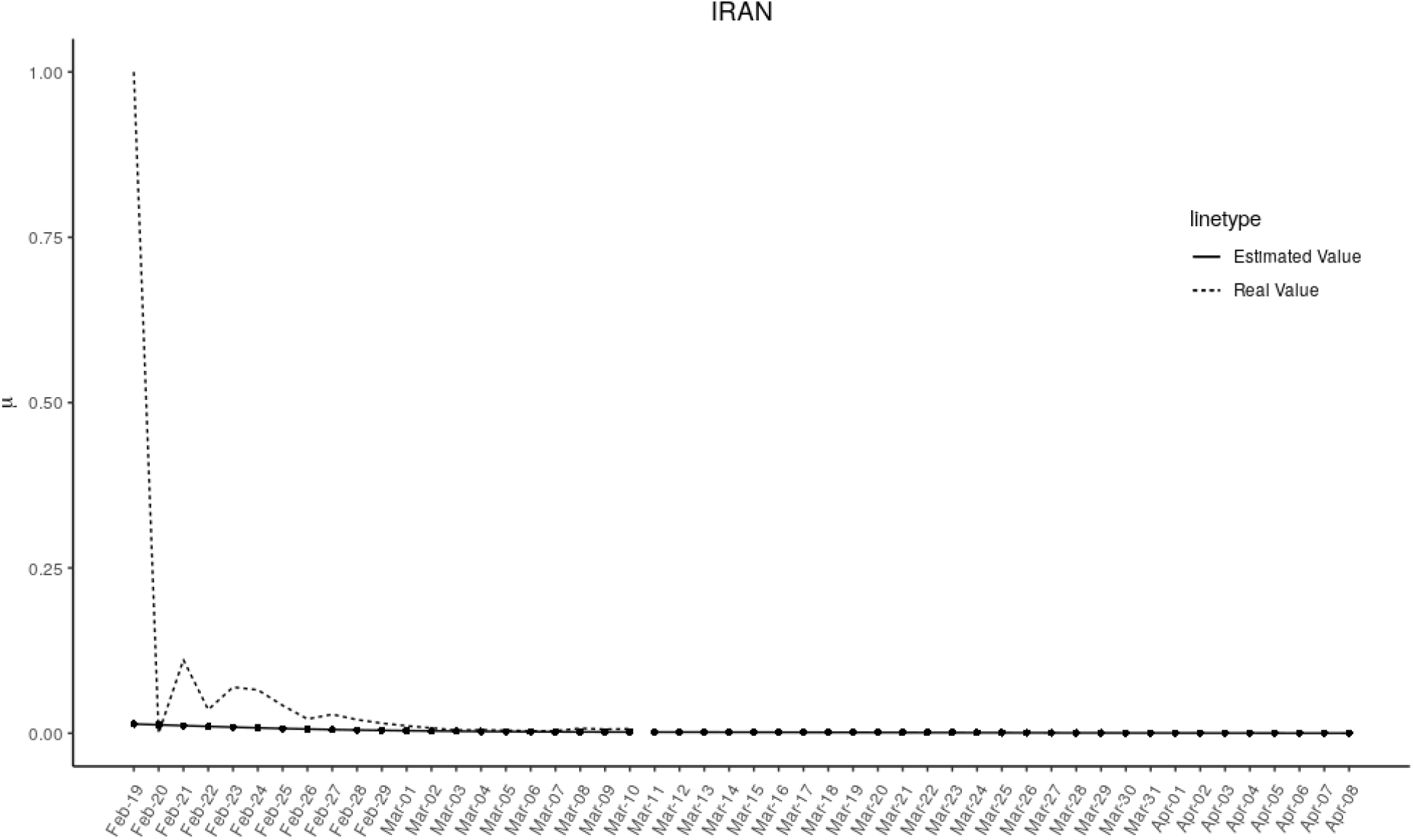
Time-dependent death rate people in Iran with μ (t) estimated of China.

**Step3**: The conclusion of step 2 has many benefits for us because we cannot obtain the behavior of the parameters for Iran. Accordingly, we can use the Chinese parameters to predict the number of Iranian patients in the last step (Figure 8).

**Figure 8:**
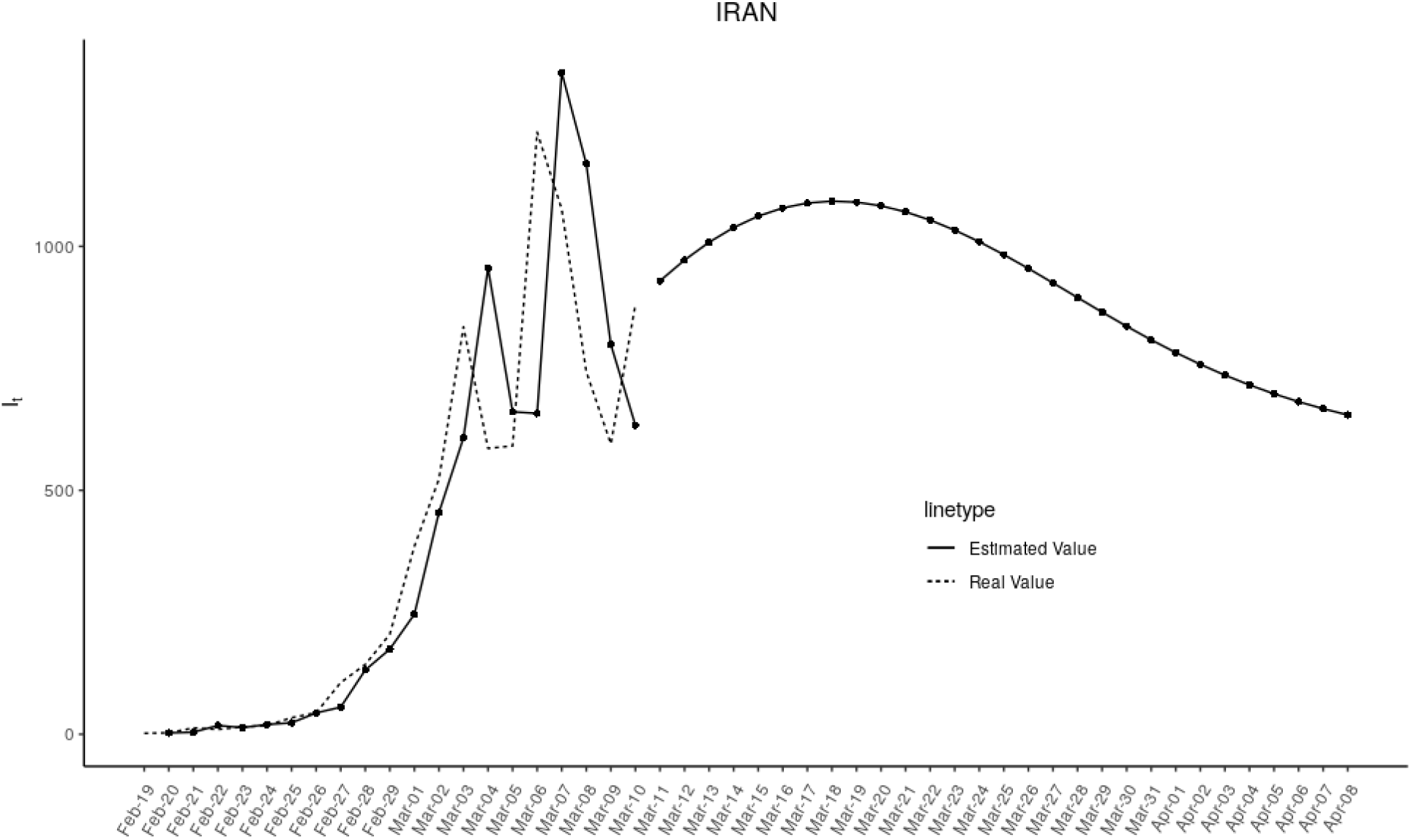
Predict the number of infected people in Iran from 03.12.2020 to 04.10.2020.

But the question remained on how to fit each parameter in China model. Many data in the environmental sciences do not fit simple linear models and are best described by “wiggly models”, also known as Generalized Additive Models (GAMs). We used GAMs to estimate β(t),(*t*),(*t*),*I*(*t*) and models fit by REML (Restricted Maximum likelihood)

One of the main reasons for using GAMs models was that we did not know the distributions of parameters, so we could not use a parametric approach like non-linear least squares (NLS).

We did all the analysis with R software. Also, mgcv package in R was used to fit generalized additive models.

## Result

The first phase (03.12.2020 - 03.15.2020): We expect daily diagnoses to increase from 1002 to 1095 until the end of the initial phase.

The third phase (03.26.2020 - 04.10.2020): In this phase, it is expected that the downward trend will continue until 04.10.2020 if the intervention measures continue at current intensity.

Assuming that the epidemic continues to develop from 03.12.2020 to 4.10.2020, 27752 people are expected to be diagnosed with COVID19 across the country. In other words, on average, 925 people with COVID 19 are expected to be infected daily in Iran. The epidemic peaks within one week (15.03.2020 to 03.21.2020) and reaches its highest point on 03.18.2020 with 1126 infected cases (Figure 8) (Table 1).

**Table 1.** Number of infected people in Iran from 03.12.2020 to 04.10.2020

| Date | I(t) | Date | I(t) |
| --- | --- | --- | --- |
| 03.12.2020 | 1002 | 03.27.2020 | 953 |
| 03.13.2020 | 1039 | 03.28.2020 | 923 |
| 03.14.2020 | 1070 | 03.29.2020 | 892 |
| 03.15.2020 | 1095 | 03.30.2020 | 862 |
| 03.16.2020 | 1112 | 03.31.2020 | 834 |
| 03.17.2020 | 1122 | 04.01.2020 | 807 |
| 03.18.2020 | 1126 | 04.02.2020 | 782 |
| 03.19.2020 | 1124 | 04.03.2020 | 760 |
| 03.20.2020 | 1116 | 04.04.2020 | 739 |
| 03.21.2020 | 1103 | 04.05.2020 | 720 |
| 03.22.2020 | 1086 | 04.06.2020 | 704 |
| 03.23.2020 | 1064 | 04.07.2020 | 688 |
| 03.24.2020 | 1040 | 04.08.2020 | 675 |
| 03.25.2020 | 1013 | 04.09.2020 | 663 |
| 03.26.2020 | 984 | 04.10.2020 | 653 |

## Discussion

Our estimation show that the COVID-19 epidemic trend in Iran will rise from March 8, 2020 and will peak during March 15 to 21, and there will be a decreasing trend from 22 to 29 March 2020. Under such circumstances, two types of hypotheses can be considered for the adequacy of the current measures and as to whether the epidemic peak will occur during March 15-21, 2020. Assuming that the current measures are inefficient and inadequate, the number of estimated cases will continue to increase by the end of March. On the other hand, the current control measures are effective and sufficient and the number of cases estimated after the epidemic peak will go down. This model may be close to reality if intervention measures continue at the current intensity (8).

At the time of the outbreak of COVID-19, the best and most urgent steps must be taken to overcome the coronavirus epidemic. The fight against coronavirus infection should be an emergency. It can only be overcome with the active cooperation of various businesses and professions such as the medical industry, transportation, government, manufacturers of technology products, etc. (9).

Several essential tasks must be done while controlling the outbreak. 1. Controlling the virus source. In the face of the virus, the most effective strategy is to keep any suspected infected patient in a relatively confined space (hospital or home) to prevent the virus from spreading. Verified patients should be treated at the hospital immediately. 2. Controlling the virus spread. Restrictive measures must be taken against the general population or the population that may spread the disease to prevent a major epidemic from continuing. 3. Virus tracking. In addition, the main source of the virus must be traced to understand the origin of the virus and effective measures must be taken to completely eliminate the source of the virus (9-11).

For an entire contaminated or potentially contaminated city, continuous air detection must be carried out to effectively detect or trace the virus in the air, and a mask must be worn by any citizen, especially in public places. New mobile hospitals should be built that can handle suspected cases in a centralized manner. This is essential to alleviate the pressure of treating the masses of infected patients in large hospitals. For example, during this outbreak, the Huoshenshan Hospital in Wuhan lasted only 10 days to be built for people infected with coronavirus. During the whole process, the epidemic situation should be reported in a timely and transparent manner. It also prevents unexpected public pressure and reduces unnecessary mental health problems.

Artificial intelligence technology can play a key role in almost every aspect including traffic management, infection detection, logistics supply chain, etc. This is a very important feature of a modern data-driven smart city. If the status of each citizen is listed, all can be accurately tracked and any population can be accommodated. Therefore, the flow of population can be controlled in a more orderly manner. Artificial intelligence technologies can be used to employ smart devices to support diagnosis and treatment, and can be used in telecommunications, online training and intelligent manufacturing to ensure minimal disruption to people’s lives. Some hospitals use smart systems. Train stations can install powerful thermal imagers to measure the body temperature of passengers. Overall, efficiency and speed are very important in the control process, and conflicting research needs to be conducted (9-11).

In response to the outbreak, China expanded its day-to-day contact with the WHO, regardless of economic disadvantages, and developed comprehensive multifaceted approaches to tackle the virus and prevent further exposure to the virus by taking rigorous and unprecedented measures arising from a sense of responsibility to citizens. It also disseminated its epidemic information openly, transparently, responsibly and in a timely manner to all other countries and international organizations (8).

There are difficult decisions for governments to make. How people respond to recommendations on preventing transmission is more important than government measures. The government’s strategy of communicating with the public to inform them about infection prevention is also crucial. Extra support for managing the recession is also important. However, it is not clear that other countries can implement the measures that the Chinese eventually adopted to reduce and control the incidence in the future.

Other countries can adopt only certain elements of China’s strategy, including suspension of public transportation, closure of recreational areas and a ban on public assemblies that have been the most effective measures to reduce the epidemic in China. Hospitals can increase their storage of medical equipment and number of beds, and the government can emphasize the importance of hand washing and staying home if ill (12).

Extensive capacity is needed for such a level of medical care, which is a health emergency and is particularly challenging. We hope that the analytical results of this study will help to clarify important aspects of the outbreak so that the spread of coronavirus in different locations can be minimized in the shortest possible time.

One of the limitations of this study is the uncertainty of data on the first days of the epidemic in Iran. The distribution of data in the first week of the epidemic was such that the fitting of each parameter of the model with Iranian data through NLS or polynomial regression models posed problems so that we could not identify a better model with AIC or BIC criteria

## Conclusion

The most important point is to emphasize the timing of the epidemic peak, hospital readiness, government measures and public readiness to reduce social contact.

## Data Availability

We statement regarding the availability of all data referred to in the manuscript and note links below.

## Contributors

Data analysis was led by BZ and AR who programmed the model with help from GM and MAM planned the inference framework. Data provided the data from online sources. MAR, BZ, GM and MAM wrote the paper. All authors interpreted the findings, contributed to writing the manuscript, and approved the final version for publication.

